# A three-dose MVA-BN mpox vaccination series improves the quality of anti-monkeypox virus immunity

**DOI:** 10.1101/2025.10.31.25339252

**Authors:** Aaron L. Oom, Kesi K. Wilson, Stephanie Rettig, Michael Tuen, Marie I. Samanovic, Angelica C. Kottkamp, Ramin Sedaghat Herati, Ralf Duerr, Mark J. Mulligan, the NYC OSMI Study Group

## Abstract

The 2022 global outbreak of clade IIb mpox represented a turning point in public health’s handling of poxviruses. The primary vaccine available for prevention of mpox is modified vaccinia Ankara from Bavarian-Nordic (MVA-BN). We previously reported a nondurable and low avidity antibody response against mpox elicited by MVA-BN. In this study, we expanded upon this knowledge by employing a microneutralization assay to measure monkeypox virus (MPXV) neutralizing titers and a multiplexed immunoassay to assess IgG titers and avidity against eight MPXV antigens and two vaccinia antigens. Through a machine learning analysis, we uncovered that MVA-BN vaccinees without prior smallpox vaccination largely return to a baseline seroprofile within a year of immunization. Notably, we identified a discrete population within this group that mounted a robust neutralizing antibody response associated with a longer dosing interval during the MVA-BN primary series. Furthermore, we found that boosting with a third dose of MVA-BN increases IgG avidity against certain MPXV antigens, as part of a booster-specific seroprofile. These findings provide critical insights into optimizing immune responses through MVA-BN boosters, presenting a promising approach to addressing the limited MPXV-specific immunity observed following the primary series.

## Introduction

The 2022 global outbreak of clade IIb mpox represented a major shift in poxvirus public health causing nearly 110,000 cases since May 2022 in non-endemic countries (1). While cases from this outbreak have since decreased from a peak of over 30,000 cases during August 2022, World Health Organization reporting demonstrates an ongoing threat from clade IIb monkeypox virus (MPXV) with cases ranging from approximately 500 to 1,500 per month across non-African nations for the past year (1). In addition to this clade IIb outbreak, there is a continuing outbreak of the more lethal clade I MPXV centered in the Democratic Republic of Congo which has resulted in almost 41,000 confirmed cases in the last year alone (1). It is important to note that the 2022 clade IIb outbreak has been genetically linked to a 2017 clade II outbreak that began in Nigeria, with host-driven APOBEC3 mutations rapidly generating multiple lineages of clade IIb virus before spreading beyond Nigeria (2,3). These findings raise concerns that the ongoing clade I outbreak could pose a similar threat of expanding globally if not properly controlled.

Current public health measures for mpox primarily rely on the third-generation orthopoxvirus (OPXV) vaccine, modified vaccinia Ankara from Bavarian-Nordic (MVA-BN, also known as JYNNEOS, Imvamune, or Imvanex). MVA-BN is a two-dose vaccine administered subcutaneously (SC) with a 28-day dosing interval. Vaccine shortages during the 2022 mpox outbreak led to authorization of longer dosing intervals as well as intradermal (ID) administration of a smaller vaccine dose. MVA-BN uses the heavily attenuated vaccinia virus (VACV)-derived MVA for its improved safety and tolerability profile over earlier generations of OPXV vaccines, particularly in immunocompromised individuals, due to its non-replicating nature (4). The earlier generations of OPXV vaccines relied on various strains of replication-competent VACV, such as New York City Board of Health, Lister, and TianTan (5), and were capable of generating durable immunity, particularly if administered in childhood (6–9). However, there is a growing body of evidence from our group and others indicating that MPXV-specific immunity induced by MVA-BN is limited in previously immunologically naïve humans (10–18). We previously showed in the New York City Observational Study of Mpox Immunity cohort (NYC OSMI) that MVA-BN generates a low avidity, nondurable MPXV-specific antibody response in individuals without prior smallpox vaccination (naïve vaccinees) (10). We also noted that an increased interval between the two doses of MVA-BN increased peak MPXV-neutralizing titers in these naïve vaccinees.

In light of these findings, there has been a growing interest in the question of protection against mpox following antibody waning (19,20). Public health data indicate that breakthrough cases of mpox generally have less severe symptoms (21–24). However, there are a number of questions that remain such as: transmissibility of breakthrough cases; impact on quarantine guidelines for these cases; and whether booster doses may further reduce the incidence of breakthrough cases. Work in the field has demonstrated that a third-dose MVA-BN booster results in higher peak OPXV neutralizing titers as compared to the peak after the standard two-dose series (4,11,25). Studies have also shown significantly slower antibody decay (4,25,26). The underlying immunological mechanisms of these improvements are unknown.

To explore these questions around boosting, we have developed a machine learning approach built on an expanded multiplexed immunoassay panel of MPXV and VACV proteins for binding studies as well as an updated MPXV microneutralization assay. With these methods we have characterized a representative subset of our NYC OSMI cohort out to two years post-vaccination and identified a distinct subgroup of naïve MVA-BN vaccinees with higher peak antibody responses, likely due to longer dosing intervals. We additionally analyzed a set of individuals who received a third-dose booster of MVA-BN, observing increased IgG avidity to certain OPXV proteins as well as the generation of a booster-specific seroprofile. Taken together, these findings indicate third-dose MVA-BN boosters as a potential strategy to overcome poor MPXV-specific immunity following the standard two-dose MVA-BN vaccine series, thereby offering critical insights for improving mpox immunity and broader poxvirus prevention efforts.

## Results

### Low avidity binding is a conserved feature of antibody responses induced by MVA-BN

For this study, we analyzed a representative subset of the NYC OSMI cohort for serological profiling. The demographics of this subset can be found in Table 1. For participants with HIV, the average CD4+ T cell counts were 671 (IQR: 466 to 883) and 846 (IQR: 557 to 1055) cells/mm^3^ for the experienced and naïve groups, respectively. Participants may enter the NYC OSMI cohort at any study visit. For this analysis, we included data from multiple study visits (Figure 1A): V1 corresponds to the baseline visit prior to vaccination, V2 occurs approximately 1 month after dose one, and V3 through V6 represent follow-up visits at ~3 weeks, ~3 months, ~9-10 months, and ~21-22 months after dose two, respectively. Only participants with at least three consecutive study visits were included. We first performed a clade IIb MPXV microneutralization assay with 5% guinea pig serum as a source of complement (27). Our findings revealed comparable peak neutralizing titers were similar between vaccinees with prior smallpox vaccination (experienced) and those without (V3 in Figure 1B), as previously observed. We also noted that there was no difference between MPXV neutralizing titers at 1 year and 2 years post-symptom onset in a group of MPXV-convalescent participants, suggesting relatively stable antibody kinetics in this group, in line with a recent report (18); convalescent participant demographics can be found in Supplementary Table S1.

**Table 1:** Demographics of NYC OSMI participant subsets used for this study.

|  | Total MVA-BN-vaccinated Participants |  | Prior Smallpox Vaccination (Experienced) |  | No Prior Smallpox Vaccination (Naïve) |  |
| --- | --- | --- | --- | --- | --- | --- |
|  | No. | % | No. | % | No. | % |
| **Enrolled Participants** | 69 |  | 30 | 43.5 | 39 | 56.5 |
| **Gender** |  |  |  |  |  |  |
| Male | 55 | 79.7 | 28 | 93.3 | 27 | 69.2 |
| Female | 10 | 14.5 | 2 | 6.7 | 8 | 20.5 |
| Non-binary/Gender Fluid/Queer | 4 | 5.8 | 0 | 0.0 | 4 | 10.3 |
| **Age** |  |  |  |  |  |  |
| Median | 46 |  | 63 |  | 33 |  |
| Range | 21 to 75 |  | 46 to 75 |  | 21 to 50 |  |
| **Race** |  |  |  |  |  |  |
| White | 48 | 69.6 | 23 | 76.7 | 25 | 64.1 |
| Black or African American | 3 | 4.3 | 3 | 10.0 | 0 | 0.0 |
| Asian | 7 | 10.1 | 2 | 6.7 | 5 | 12.8 |
| Other | 11 | 15.9 | 2 | 6.7 | 9 | 23.1 |
| **Ethnicity** |  |  |  |  |  |  |
| Non-Hispanic | 51 | 73.9 | 23 | 76.7 | 28 | 71.8 |
| Hispanic | 17 | 24.6 | 6 | 20.0 | 11 | 28.2 |
| Other | 1 | 1.4 | 1 | 3.3 | 0 | 0.0 |
| **LGBTQ+** |  |  |  |  |  |  |
| Yes | 60 | 87.0 | 29 | 96.7 | 31 | 79.5 |
| No | 9 | 13.0 | 1 | 3.3 | 8 | 20.5 |
| **Administration Route** |  |  |  |  |  |  |
| SC-SC | 16 | 23.2 | 6 | 20.0 | 10 | 25.6 |
| ID-ID | 13 | 18.8 | 7 | 23.3 | 6 | 15.4 |
| ID-SC | 9 | 13.0 | 5 | 16.7 | 4 | 10.3 |
| SC-ID | 31 | 44.9 | 12 | 40.0 | 19 | 48.7 |
| **HIV Status** |  |  |  |  |  |  |
| People with HIV | 19 | 27.5 | 13 | 43.3 | 6 | 15.4 |
| HIV-uninfected | 50 | 72.5 | 17 | 56.7 | 33 | 84.6 |

**Figure 1:**
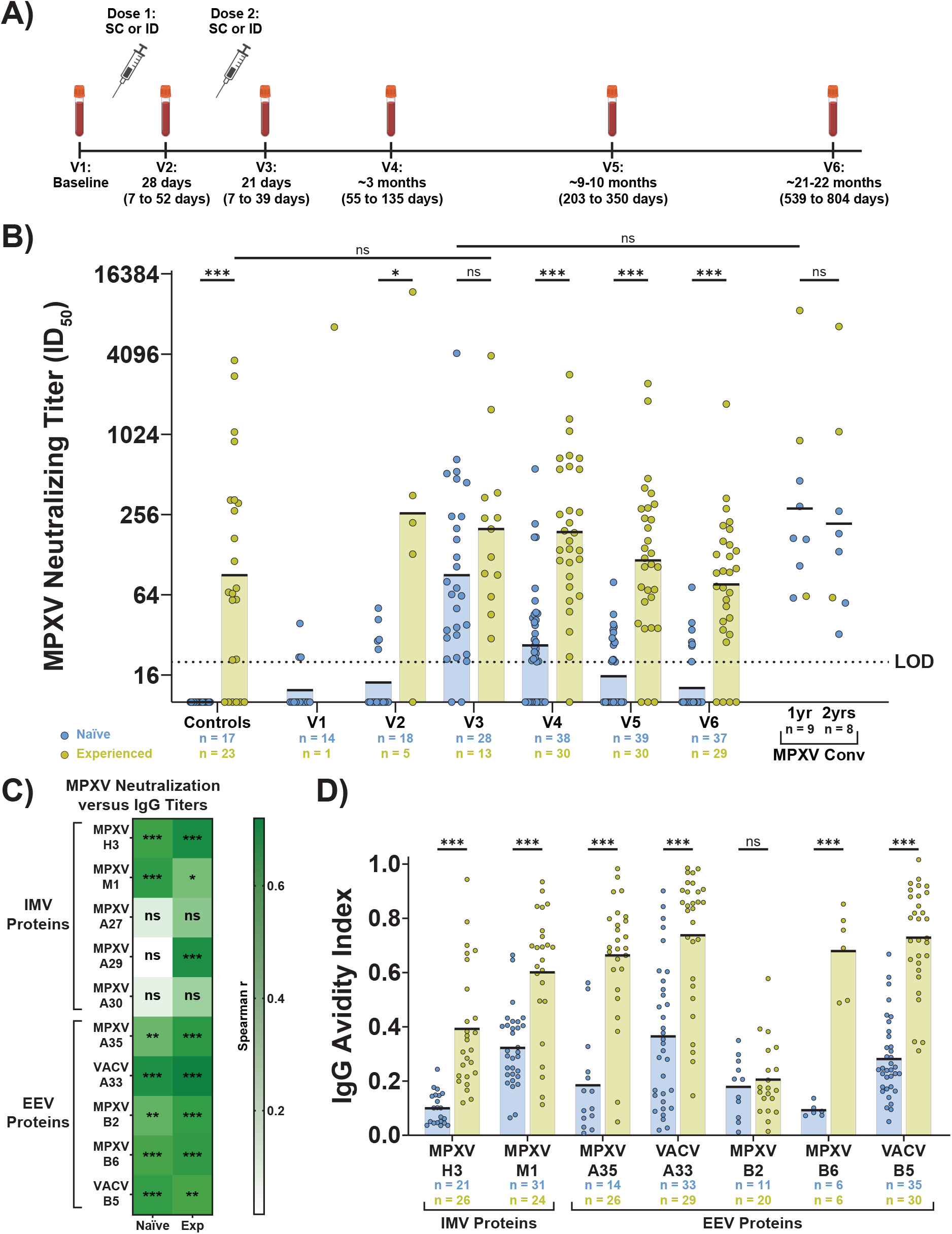
MVA-BN induces a nondurable and low avidity MPXV-specific antibody response in previously naïve vaccinees. A) Study schedule with median visit times. Ranges of visit times included in each visit are in parentheses. All times are in relation to the previous vaccine dose. Image generated with BioRender. B) Neutralizing titers against clade IIb MPXV as measured by microneutralization assay with 5% guinea pig serum. Mpox convalescent samples are from the indicated time point post-symptom onset. Naïve and experienced controls were collected pre-2022. Black bars represent the geometric mean. Samples with ID_50_ values below the limit of detection (LOD) are assigned a value of 10, or ½ the LOD. C) Spearman correlation of IgG for the indicated protein versus MPXV neutralizing titers. Samples are from V4. D) Avidity index for IgG that correlated with MPXV neutralization in both naïve and experienced vaccinees in C. Only avidity measurements from samples with above background binding are included. Black bars represent the mean. Statistics for panel B were performed using the Kruskal-Wallis test with Dunn’s method for multiple comparisons and for panel D were performed using ordinary one-way ANOVA with Šídák’s multiple comparisons test. MPXV Conv, mpox convalescent patients; IMV, intracellular mature virion; EEV, extracellular enveloped virion.

In our previous work, we focused on just two MPXV proteins, H3 and A35, for IgG binding and avidity assays. Here we expanded our panel to include a broader set of attachment and fusion proteins (proteins in bold in Table 2). This protein panel was selected to include a mix of both intracellular mature virion (IMV) and extracellular enveloped virion (EEV) proteins as both viral forms are important immune targets in infection and disease control. Using the Spearman correlation test, we identified five MPXV proteins and two VACV proteins for which IgG binding correlated with MPXV neutralization in naïve and experienced vaccinees at V4 (Figure 1C): MPXV H3, M1, A35, B2, and B6, and VACV A33 and B5, the homologs of MPXV A35 and B6, respectively. MPXV A29 was an additional correlate for experienced vaccinees only. Spearman r values were comparable between IMV and EEV proteins for both naïve and experienced vaccinees. To measure IgG avidity, we used a chaotropic immunoassay that includes a wash step with 2M ammonium thiocyanate. We found that IgG against six of the seven target proteins had lower avidity among the naïve vaccinees as compared to the experienced vaccinees (Figure 1D). For MPXV B2 there was equally low avidity in the naïve and experienced vaccinees. Taken together these data indicate that low IgG avidity against the viral proteins that correlate with MPXV neutralization is a conserved feature of *de novo* antibody responses generated by MVA-BN.

**Table 2:** Panel of proteins used for multiplexed immunoassay.

| **MPXV Protein** | **VACV Copenhagen Homolog** | **Function/Location** | **References** |
| --- | --- | --- | --- |
| **H3** | H3 | IMV protein, binding | Ref. (44,45) |
| **M1** | L1 | IMV protein, member of entry-fusion complex (EFC) | Ref. (46–48) |
| **A27** | A26 | IMV protein, pH-dependent fusion suppressor | Ref. (49) |
| **A29** | A27 | IMV protein, binding | Ref. (48,50) |
| **A30** | A28 | IMV protein, EFC member | Ref. (51) |
| **A35** | **A33** | EEV protein, envelope glycoprotein | Ref. (38,39,48) |
| **B2** | A56 | EEV protein | Ref. (52) |
| **B6** | **B5** | EEV protein, actin tail formation | Ref. (44,48) |

Ten proteins in **bold** are included in this study.

### MVA-BN vaccinees return to a baseline-like seroprofile by one year post-vaccination

To identify broader trends across the multiple serologic dimensions measured, we applied machine learning analyses. We included neutralizing titers as well as IgG binding titers and avidity measurements for each of the eight viral protein correlates of neutralization, along with data for MPXV A27, as binding titers against A27 demonstrate clear delineation between replicating and non-replicating OPXV exposure (Supplementary Figure S1). Data for each sample were reduced to two dimensions using principal component analysis (PCA) then clustered using K means clustering. The optimal number of clusters was determined based on silhouette scores (see Methods). The PCA plot revealed clear segregation of naïve versus experienced visits (Figure 2A, O’s versus X’s on the plot, respectively), with IgG avidity emerging as the major driver in this separation, as shown in the biplot (Figure 2B). This underscores the importance of IgG quality in our understanding of MPXV seroprofiles.

**Figure 2:**
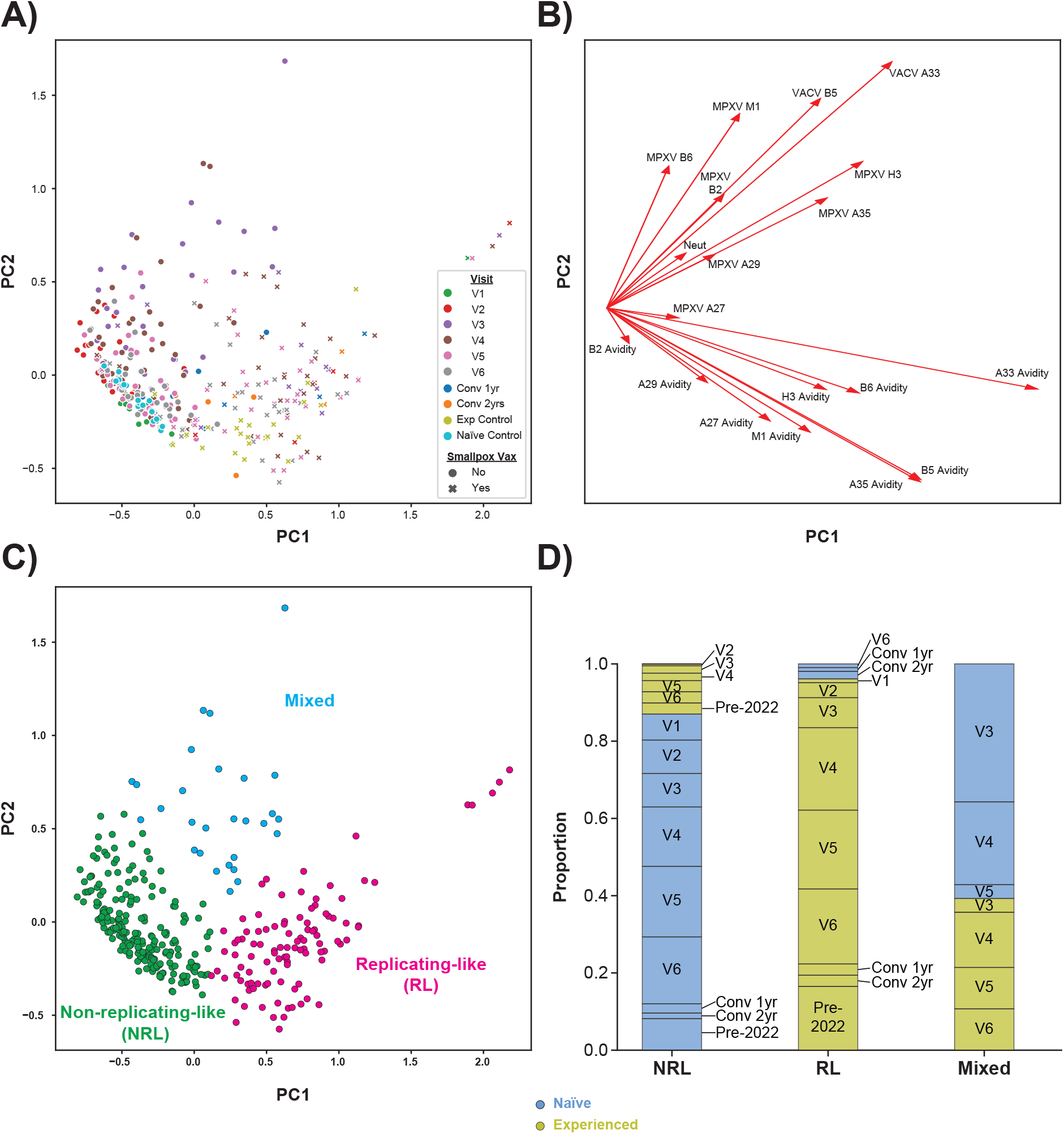
MPXV seroprofiles are driven by avidity measurements and reveal a distinct population of naïve vaccinees. A) Principal component analysis (PCA) plot of full cohort. B) Biplot of principal component loadings. C) Labeled PCA plot from A following K means clustering. D) Proportional composition of clusters from C.

Following K means clustering, we observed three clusters that corresponded to individuals with a non-replicating first OPXV vaccination (i.e., MVA-BN), replicating first OPXV vaccination (i.e., VACV), and a mixed population (Figure 2C). Interestingly, the “non-replicating-like” cluster (NRL, green in Figure 2C) is primarily composed of visits from naïve participants across early time points (pre-dose 2) as well as visits from the 1- and 2-year time points, V5 and V6, respectively (Figure 2D). Notably, nearly all V5 and V6 time points for naïve vaccinees clustered within the NRL group, with the exception of a single V6 sample that clustered with “replicating-like” group (RL) along the boundary. This suggests that MVA-BN vaccinees serologically return to a baseline-like state by one year post-vaccination. A similar finding was reported recently for a serology-based diagnostic approach that saw poor discrimination of vaccinated individuals from negative controls at seven months and later after vaccination (28).

### MPXV seroprofiles are consistent across MVA-BN cohorts

To validate the seroprofiles found for the NYC OSMI cohort, we tested 24 samples from a validation cohort (Crandell et al. (12)). The validation samples include 9 smallpox vaccinees (Dryvax group or VACV-only), 8 naïve MVA-BN vaccinees, and 7 experienced MVA-BN vaccinees. Individual sample details are reported in Table 3 and an overlay of the samples on the NYC OSMI PCA plot can be found in Supplementary Figure S2. The naïve MVA-BN vaccinees from this cohort were all classified within the NRL group, while experienced MVA-BN vaccinees and VACV-only vaccinees split between the RL and NRL groups. This latter point is similar to what was seen with NYC OSMI samples with a proportion of VACV-vaccinated individuals clustering with the NRL group (Figure 2D). This overlap reinforces the validity of the seroprofiles, demonstrating their consistency across independent cohorts of MVA-BN and VACV vaccinees.

**Table 3:** Classification of samples from Crandell et al (12).

| Subject ID | Age Range | Sex | Vaccine History | Dosing Interval | DPV2 | Assigned Group | Assignment Probability |
| --- | --- | --- | --- | --- | --- | --- | --- |
| Yale 01 | 50-59 | Female | Dryvax |  |  | NRL | 1.000 |
| Yale 02 | 50-59 | Male | Dryvax |  |  | NRL | 1.000 |
| Yale 03 | 50-59 | Female | Dryvax |  |  | RL | 1.000 |
| Yale 04 | 50-59 | Female | Dryvax |  |  | NRL | 1.000 |
| Yale 05 | 50-59 | Male | Dryvax |  |  | RL | 1.000 |
| Yale 06 | 30-39 | Female | Dryvax |  |  | RL | 1.000 |
| Yale 07 | 50-59 | Female | Dryvax |  |  | RL | 0.996 |
| Yale 08 | 50-59 | Female | Dryvax |  |  | NRL | 1.000 |
| Yale 09 | 40-49 | Male | Dryvax |  |  | RL | 1.000 |
| Yale 10 | 60-69 | Male | Dryvax +MVA-BN | 30 | 8 | RL | 0.932 |
| Yale 11 | 60-69 | Male | Dryvax +MVA-BN | 30 | 60 | RL | 0.816 |
| Yale 12 | 50-59 | Male | Dryvax +MVA-BN | 32 | 180 | RL | 1.000 |
| Yale 13 | 40-49 | Male | Dryvax +MVA-BN | 28 | 240 | NRL | 1.000 |
| Yale 14 | 60-69 | Male | Dryvax +MVA-BN | 28 | 270 | RL | 1.000 |
| Yale 15 | 40-49 | Male | Dryvax +MVA-BN | 30 | 11 | NRL | 1.000 |
| Yale 16 | 50-59 | Male | Dryvax +MVA-BN | 30 | 270 | RL | 1.000 |
| Yale 17 | 30-39 | Male | MVA-BN | 33 | 64 | NRL | 1.000 |
| Yale 18 | 30-39 | Male | MVA-BN | 42 | 149 | NRL | 1.000 |
| Yale 19 | 20-29 | Male | MVA-BN | 36 | 280 | NRL | 1.000 |
| Yale 20 | 30-39 | Male | MVA-BN | 30 | 67 | NRL | 0.996 |
| Yale 21 | 20-29 | Male | MVA-BN | 28 | 298 | NRL | 1.000 |
| Yale 22 | 30-39 | Male | MVA-BN | 30 | 238 | NRL | 1.000 |
| Yale 23 | 20-29 | Female | MVA-BN | 28 | 99 | NRL | 1.000 |
| Yale 24 | 20-29 | Male | MVA-BN | 28 | 307 | NRL | 1.000 |

### Robust Responders are characterized by higher peak neutralizing titers associated with longer dosing intervals

Following seroprofile validation, we further examined the composition of the three clusters. Among the naïve participants, we identified a subset of V3 and V4 sera that were within the mixed population (blue in Figure 2C) as opposed to the NRL group. These samples accounted for ~1/3 of the naïve vaccinees at V3. We found higher MPXV neutralization among this subset as compared to participants in the NRL cluster at V3 (Figure 3A). Thus, we deemed these individuals Robust Responders, while those with V3 in the NRL cluster were labeled Standard Responders. To determine whether being a Robust Responder provided long-term benefits, we compared the MPXV neutralizing titers at V4 (Figure 3B), V5 (Figure 3C), and V6 (Figure 3D). This advantage in neutralizing titers persists at V4. But, in line with the observation that most V5 and V6 samples are found within the NRL cluster, there was no difference between Robust and Standard Responders at either V5 or V6 for neutralization. Additionally, we saw no difference in IgG avidity between Robust and Standard Responders at V4 (Figure 3E).

**Figure 3:**
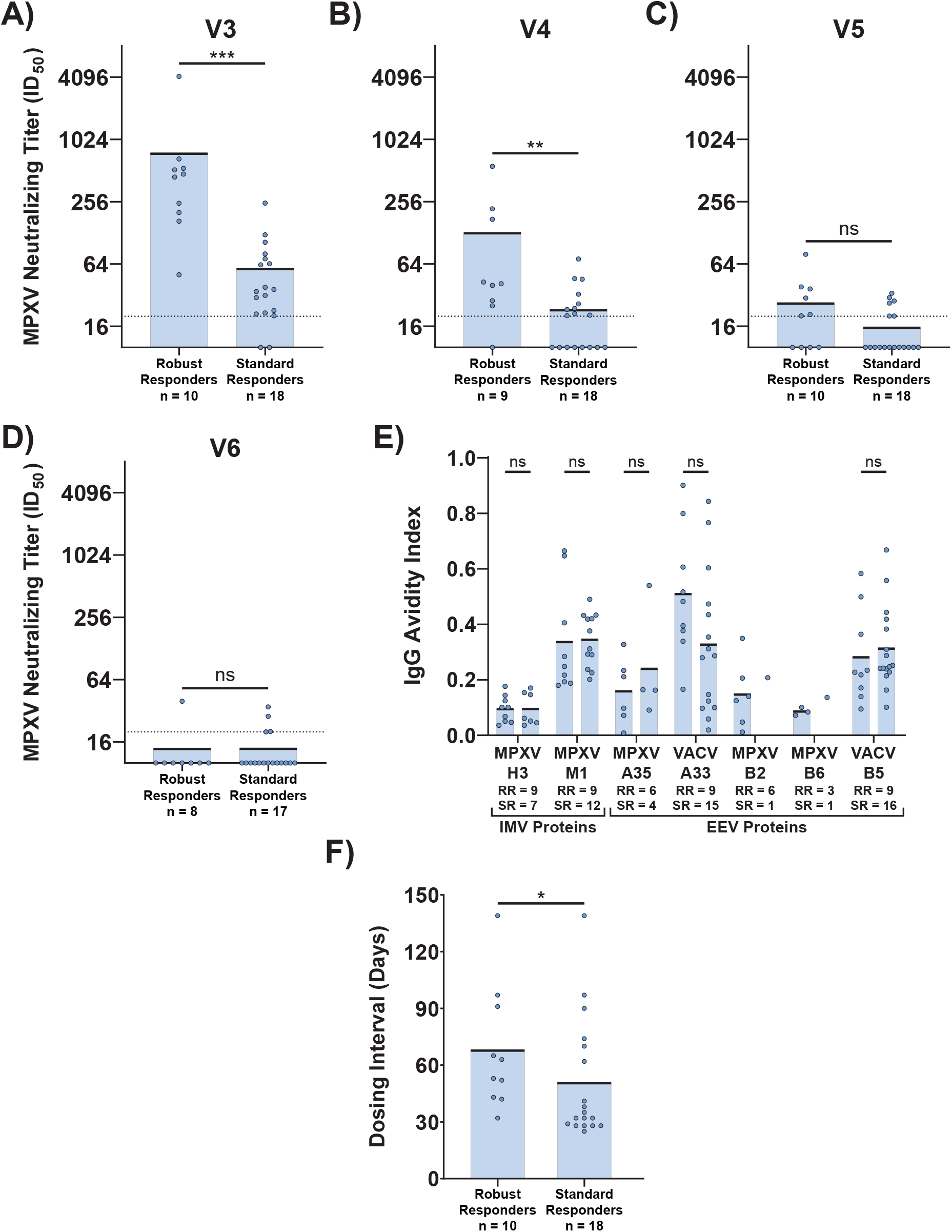
Robust Responders have a higher peak MPXV neutralizing titer and longer dosing interval. A-D) Comparison of MPXV neutralizing titer as assayed in Figure 1 of Robust versus Standard Responders for V3 (A), V4 (B), V5 (C), and V6 (D). E) Comparison of IgG avidity for Robust versus Standard Responders. F) Comparison of dosing interval for Robust versus Standard Responders. Black bars on all panels represent the mean. Statistics for panels A through D and F were performed using the Mann-Whitney test and for panel E were performed using ordinary one-way ANOVA with Šídák’s multiple comparisons test.

To determine features distinguishing Robust Responders from Standard Responders, we next compared demographics and clinical variables (Table 4). We found no differences in age, gender, HIV status, or route of vaccine administration between the two groups. However, there was a difference in the time between doses, or dosing interval (Figure 3F). This is in line with our previous finding that longer dosing intervals correlated with higher peak MPXV neutralizing titers (10). Taken together, these data indicate that longer dosing intervals may enhance the magnitude, but not the quality, of antibody response elicited by MVA-BN vaccination.

**Table 4:** Comparison of Robust versus Standard Responders.

|  | **Robust Responders** |  | **Standard Responders** |  | **P-value** |
| --- | --- | --- | --- | --- | --- |
|  | **No.** | **%** | **No.** | **%** |  |
| **Participants** | 10 |  | 18 |  |  |
| **Gender** |  |  |  |  |  |
| Male | 7 | 70.0 | 13 | 72.2 | >0.999<br>(Fisher's Exact) |
| Female | 2 | 20.0 | 4 | 22.2 |  |
| Non-binary/Gender Fluid/Queer | 1 | 10.0 | 1 | 5.6 |  |
| **Age** |  |  |  |  |  |
| Median | 30 |  | 34 |  | 0.141<br>(Mann-Whitney) |
| Range | 24 to 41 |  | 23 to 50 |  |  |
| **Dosing Interval** |  |  |  |  |  |
| Median | 58 |  | 34 |  | **0.046**<br>(Mann-Whitney) |
| IQR | 43-93 |  | 28-71 |  |  |
| **Administration Route** |  |  |  |  |  |
| SC-SC | 3 | 30.0 | 7 | 38.9 | 0.367<br>(Fisher's Exact) |
| ID-ID | 5 | 50.0 | 3 | 16.7 |  |
| ID-SC | 1 | 10.0 | 3 | 16.7 |  |
| SC-ID | 1 | 10.0 | 5 | 27.8 |  |
| **HIV Status** |  |  |  |  |  |
| People with HIV | 9 | 90.0 | 15 | 83.3 | >0.999<br>(Fisher's Exact) |
| HIV-uninfected | 1 | 10.0 | 3 | 16.7 |  |

Statistical significance test is indicated below the respective p-value.

### MVA-BN third-dose boosters improve IgG avidity for select MPXV proteins

In light of reports demonstrating improved neutralizing titers following MVA-BN third-dose boosters, we hypothesized that a third dose of non-replicating MVA-BN would also improve antibody quality, i.e., avidity. To test this, we assayed samples from four NYC OSMI participants at risk of occupational OPXV exposure who received a third-dose MVA-BN booster (Table 5). Two were Robust Responders following two MVA-BN doses (subjects Booster 1 and 4) while two were Standard Responders (Booster 2 and 3), offering an opportunity to determine if that status impacted booster responses. All four participants demonstrated clear responses to the third dose, including Booster 3 who was a “non-responder” to the primary vaccine series (Figure 4A). While all four participants showed a higher peak neutralizing titer following dose three as compared to dose two, this difference did not reach statistical significance (Figure 4B, p = 0.1250). Notably, IgG avidity against MPXV H3 (Figure 4C) and M1 (Figure 4D), and VACV B5 (Figure 4E) was higher following boosting. However, this improvement was not seen for IgG against MPXV A35 (Figure 4F) or its VACV homolog, A33 (Figure 4G). These data indicate an improvement of anti-OPXV IgG quality (avidity) against a subset of MPXV proteins with a third-dose booster of MVA-BN.

**Table 5:** Demographics of MVA-BN booster participants.

| ID | Age Range | Gender | Dosing Interval (Dose 1-2) | Dosing Interval (Dose 2-3) | Routes of Administration | Marker | Dose 2 Group |
| --- | --- | --- | --- | --- | --- | --- | --- |
| 1 | 20-29 | Male | 42 | 725 | SC-ID-SC | ▲ | Robust Responder |
| 2 | 30-39 | Female | 28 | 771 | SC-SC-SC | ◆ | Standard Responder |
| 3 | 50-59 | Male | 28 | 989 | SC-SC-SC | ■ | Standard Responder |
| 4 | 40-49 | Female | 32 | 1024 | SC-SC-SC | ● | Robust Responder |

Indicated marker is used in Figures 4, 5, and 6. All participants are HIV negative.

**Figure 4:**
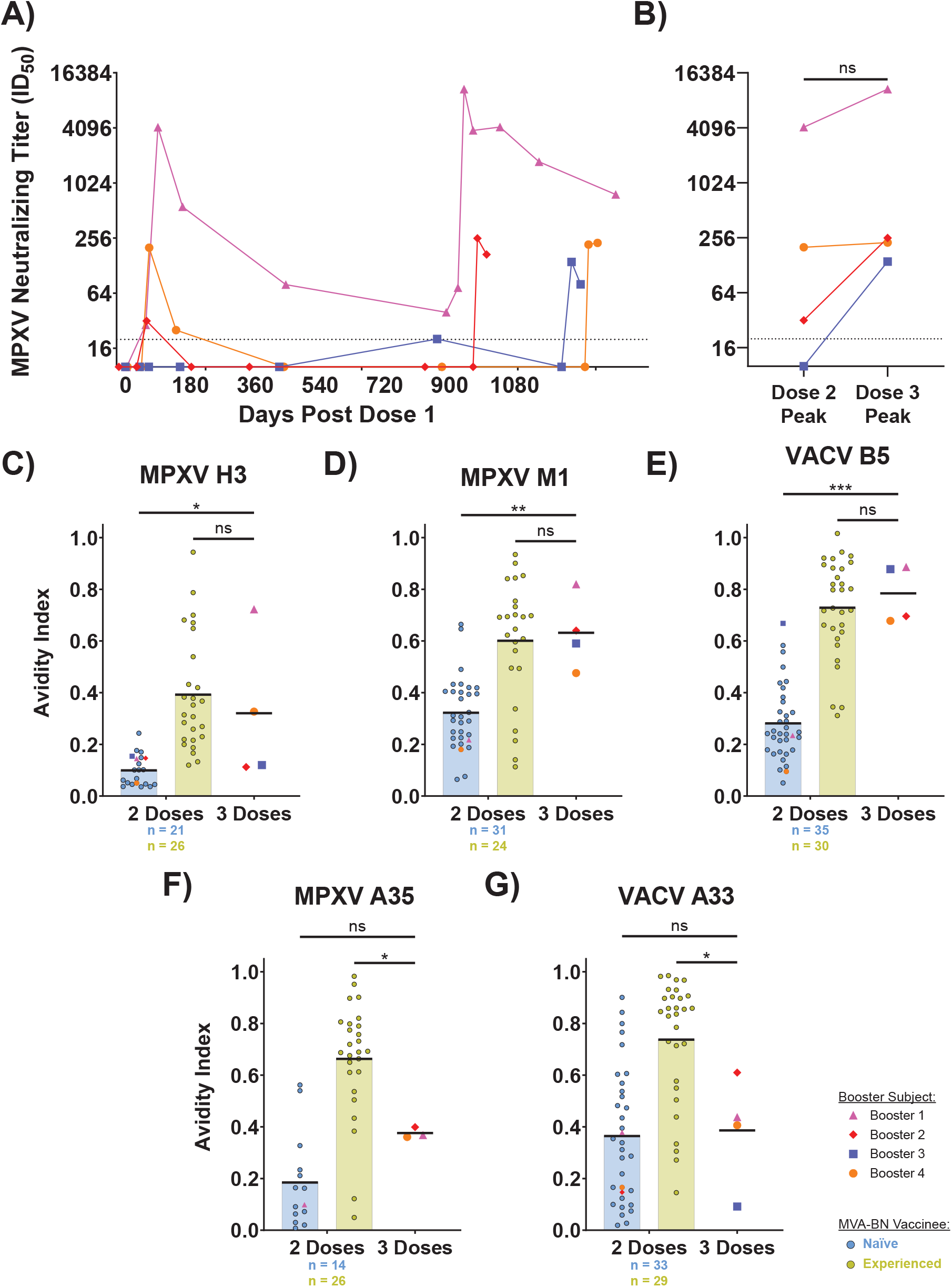
MVA-BN boosting improves IgG avidity for select OPXV-specific antibodies. A) MPXV neutralization as assayed in Figure 1 for booster subjects in Table 5. B) Comparison of peak neutralizing titers following dose two and three. C-G) Avidity index for IgG against MPXV H3 (C), MPXV M1 (D), VACV B5 (E), MPXV A35 (F), and VACV A33 (G). Avidity measurements for post-boost samples use the 1 month post-boost time point with the exception of the measurement for Booster 2 on MPXV A35, which uses the 1 week post-boost time point due to negative levels of anti-MPXV A35 binding at the 1 month time point. Measurements from Figure 1 are shown here for the two dose group. Black bars on panels C through G represent the mean. Statistics were performed using Wilcoxon matched-pairs signed rank test for panel B and ordinary one-way ANOVA with Holm-Šídák’s multiple comparisons test for panels C through G.

Our observations that a primary series of MVA-BN elicits a nondurable, low avidity antibody response raised the possibility of a primarily extrafollicular (EF) B cell response, as opposed to a germinal center (GC)-derived response. Using the automated ELISA platform ELLA, we examined serum CXCL13 levels as a correlate of GC formation (29). Within the two weeks after vaccination, we did not find any difference in serum CXCL13 following the two-dose MVA-BN primary series (Figure 5A). However, we did note a correlation between dosing interval and serum CXCL13 levels out to 14 days post-dose two in naïve vaccinees (Figure 5B). This may explain the higher neutralizing titers among vaccinees with longer dosing intervals. As we saw improvements in IgG avidity following a third-dose booster, we wondered whether this could be linked to improved GC formation. To test this, we compared levels of serum CXCL13 in our booster participants from before receipt of dose three and one week after vaccination (Figure 5C). While all participants had a rise in CXCL13, the difference did not reach statistical significance (p = 0.1228). These data suggest either limited GC formation or, alternatively, the sample size of 4 was too small to capture the difference.

**Figure 5:**
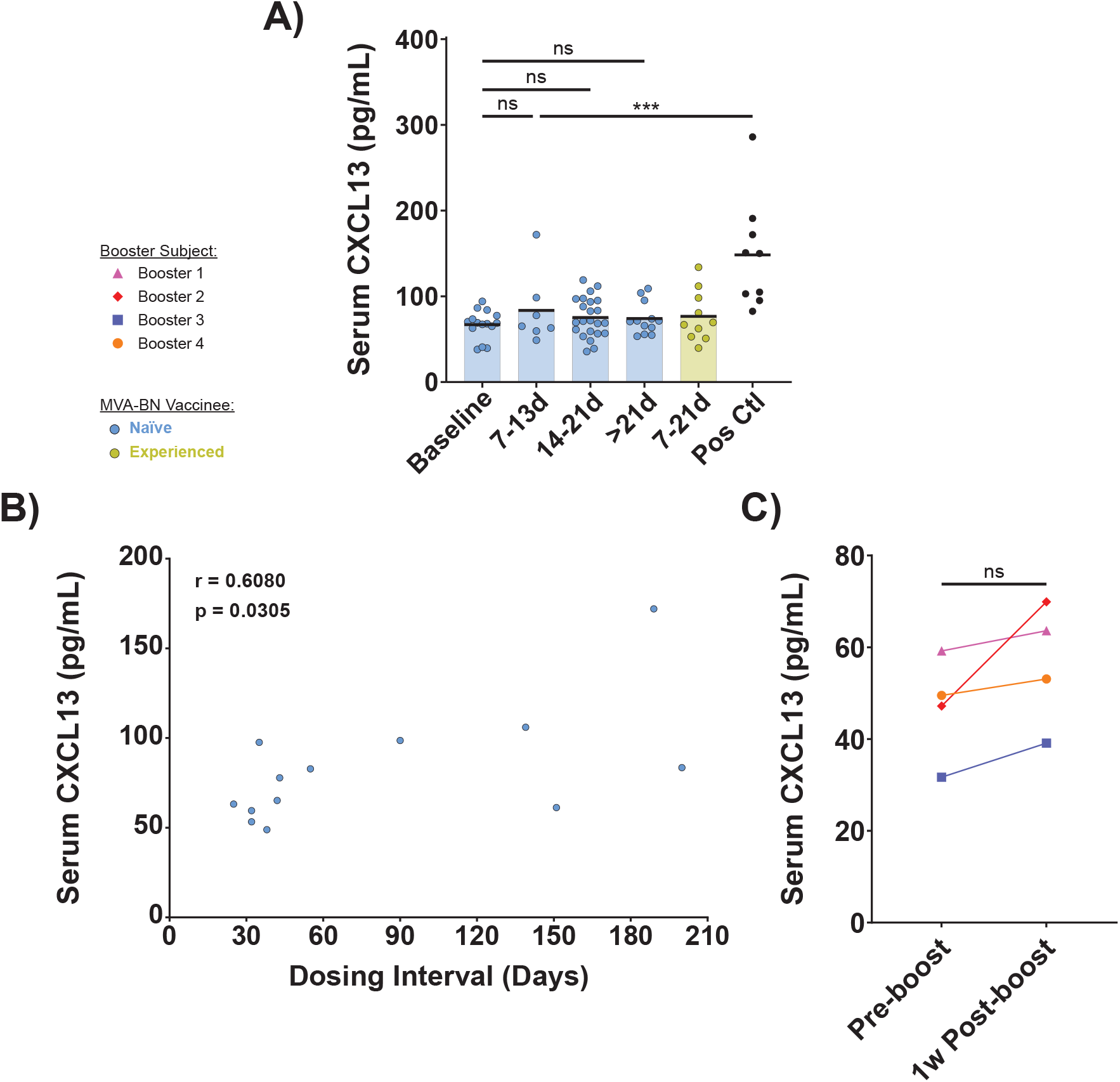
Serum CXCL13 levels indicate below threshold germinal center activity following MVA-BN vaccination. A) Serum CXCL13 levels as assayed by automated ELISA for MVA-BN vaccinees at indicated time points post-dose two. Positive control samples are from acute respiratory infection patients. Black bars represent the mean. B) Spearman correlation of dosing interval and post-dose two serum CXCL13 levels. Samples from up to 14 days post-dose two are included. C) Comparison of serum CXCL13 levels before and after third-dose MVA-BN booster. Post-boost samples were from ~1 week after vaccination. Statistics were performed using ordinary one-way ANOVA with Šídák’s multiple comparisons test for panel A and paired t test for panel C.

### MVA-BN boosting induces unique antibody profiles from VACV and primary series recipients

Finally, we wanted to understand how the seroprofiles generated by MVA-BN boosting would fit within our larger cohort. In light of the improvements in neutralizing titers and IgG avidity we hypothesized that post-booster samples may behave similarly to samples from VACV-vaccinated individuals. However, we were surprised to find that results of this analysis were more mixed. The strongest Robust Responder, subject Booster 1, was classified back into the mixed population (Figure 6A), along the border of the mixed and RL groups. Samples from Booster 2 were classified within the RL group, situated at the border of the three groups (Figure 6B), despite having previously been a Standard Responder. However, Booster 3 and 4 both remained within the NRL group following boosting (Figure 6C and D), with a clear distinction between post-dose two and post-dose three sera for Booster 4. Notably, Robust versus Standard Responder status did not offer a clear prediction of post-boosting serology behavior. Taken together, these findings indicate that MVA-BN boosting is capable of inducing improved antibody responses that are distinct from either the primary series or VACV vaccination.

**Figure 6:**
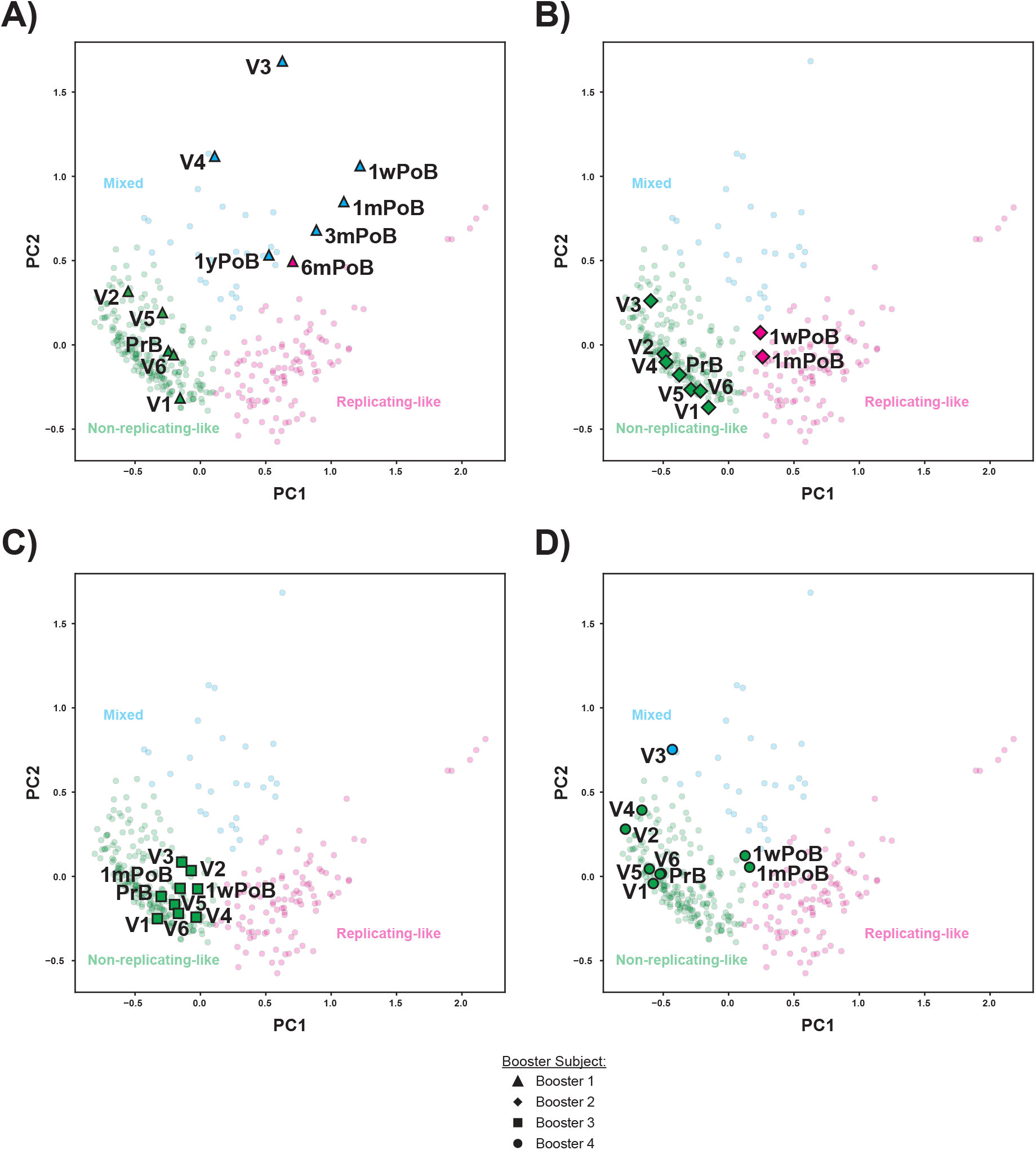
Seroprofiles following MVA-BN boosting are distinct from those after the primary series or VACV vaccination. A-D) PCA plot of all time points for subjects Booster 1 (A), Booster 2 (B), Booster 3 (C), and Booster 4 (D). Pre-boost (PrB) and post-boost (PoB) samples were classified using a bagged support vector machine classifier trained on the full NYC OSMI subset detailed in Table 1, including V1 through V6 for these participants. Post-boost samples are labeled by number of weeks (w), months (m), or years (y) following boost.

## Discussion

Building upon previous work in the field, this study provides critical new insights into the immunological landscape shaped by MVA-BN vaccination. We have demonstrated that 1) the seroprofiles of MVA-BN vaccinees return to an overall baseline state within a year of vaccination, highlighting the transient nature of the induced immunity; 2) low IgG avidity dominates the MVA-BN response; and 3) a longer dosing interval during the primary vaccination series is associated with induction of a transitory “Robust Responder” state characterized by higher peak neutralizing titers. These new results offer crucial updates to existing literature that has demonstrated a poorly durable anti-MPXV response following the MVA-BN primary series.

Our findings here also offer further clarity to previous work that demonstrated improved antibody responses following boosting (4,11,25,26). We have shown here that these booster-mediated improvements extend to antibody quality in terms of avidity. These results highlight the potential of booster doses to address the insufficient durability of protection provided by the primary series. The addition of a third dose offers an accessible and straightforward strategy for enhancing mpox immunity, particularly in settings where public health officials confirm insufficient protection from the two-dose series. However, this research adds an important dimension to our understanding of the limitations of non-replicating poxvirus vaccines, as even boosting with a third MVA-BN dose does not fully recreate the seroprofile induced by initial smallpox vaccination with replicating VACV. Further studies are needed to determine what a useful threshold of immunity looks like as it pertains to protection and will inform development of subsequent generations of improved OPXV vaccines.

In examining potential mechanisms of short-lived antibody responses following MVA-BN vaccination, we explored the hypothesis that the two-dose primary series elicits an EF maturation path for B cell development while the third dose induces a GC maturation pathway. This hypothesis comes from a model of B cell development that requires passage through the GC in order to form high affinity, long-lived plasma cells, although this remains a point of contention within the field (30). The use of serum CXCL13 as a correlate of GC formation has limitations and may be a phenomenon that requires surpassing some threshold of GC activity, which means a negative result does not strictly rule out GC formation. To this point, we previously found no significant rise in serum CXCL13 following vaccination with COVID-19 mRNA shots (31) despite clear evidence of robust GC formation in this setting (32). Further examination of this question for MVA-BN will require analysis of antigen-specific B cells to determine the degree of somatic hypermutation in these B cell receptors as well as transcriptional and epigenetic profiles.

The role of dosing intervals also emerged from this study as a critical factor in defining antibody responses. This was similarly observed during the COVID-19 pandemic, where longer dosing intervals enhanced both humoral and cellular immunity (33–35). A similar public health dilemma was faced during the 2022 clade IIb mpox outbreak leading to varying dosing intervals. The association of Robust Responder status with a longer dosing interval indicates that changes to the dosing schedule may improve antibody responses following MVA-BN vaccination, but we note the transient nature of this status at least in these data. As most of the vaccinees studied here received their second dose within three months after their first, it is possible that further extension of the dosing interval is required to achieve a prolonged benefit. Controlled clinical studies will be needed in order to answer this question.

There are several limitations to our work. While we validated the seroprofiles detailed here in another independent cohort, study of samples from further cohorts will strengthen the generalizability of these profiles. With regards to the booster studies, we have examined only a small cohort. In addition, our booster cohort comprises only HIV-negative individuals; people with HIV remain an important demographic for MVA-BN vaccination and protection from mpox. However, we believe that these data offer an important foundation for further characterizing immunity following MVA-BN boosting. Finally, humoral immunity, as studied here, is only a part of the picture when it comes to protection from infection or disease. The contributions of long-term memory B and T cells are still poorly understood in the context of MVA-BN.

Despite vaccination efforts, mpox remains an ongoing concern both in the US and abroad. While we have not seen additional surges in the US since 2022, the CDC has already recorded nearly a thousand cases of mpox in 2025, highlighting a now endemic pattern that remains far above pre-2022 levels (36). Beyond mpox, other emerging poxviruses pose a concerning future threat to public health in a global population that is now predominantly poxvirus-naïve. Serological studies like ours contribute critical pieces to the puzzle of understanding MVA-BN-induced immunity, but much work remains to address cellular immunity, antibody maturation pathways, and opportunities to optimize vaccines against orthopoxviruses. Recent work (37–42) has begun to explore these exciting questions, offering a road map for containing the specter of future orthopoxvirus outbreaks.

## Methods

### Study Design

The NYC Observational Study of Mpox Immunity is a cohort of adults with and without HIV who have been vaccinated against mpox with MVA-BN. The cohort additionally contains individuals who were previously infected with MPXV (NCT05654883, Institutional Review Board protocol 22-01338). This study has been previously described (10,43). Briefly, participants enrolled pre- or post-MVA-BN vaccination, up to one year post-first dose or mpox symptom onset. Study visits were as follows: baseline (V1), ~1 month post-dose one (V2), and ~3 weeks, ~3 months, ~9-10 months, and ~21-22 months post-dose two (V3, V4, V5, and V6, respectively). At each visit participants were sampled for serum, peripheral blood mononuclear cells, and saliva. The full cohort contains 171 adult participants, comprising 159 MVA-BN vaccinees with full vaccination history, 11 mpox convalescent patients, and 1 MVA-BN vaccinee with incomplete vaccination history. A subset of these participants was studied for this analysis; demographics can be found in Table 1 for vaccinees and Supplementary Table S1 for convalescent patients. Sex as a biological variable was accounted for where possible, however the 2022 mpox outbreak disproportionately affected men who have sex with men. As an observational study, therefore, a high proportion of participants identify as male. The general findings detailed here are likely to apply to both the male and female sex. Participant enrollment was conducted through city vaccination centers as well as by word of mouth.

Additional pre-2022 control samples, pre-NYC OSMI enrollment participant samples, and booster samples were obtained from the NYU Langone Vaccine Center Biorepository (Institutional Review Board protocol 18-02035). Study details for samples from Crandell et al. can be found at the associated reference (12).

### Blood Collection

Venous blood was collected via standard phlebotomy for NYC OSMI. Serum was collected in SST tubes (BD Biosciences), then aliquoted and frozen immediately at −80°C. Whole blood for Crandell et al. was collected in heparinised CPT vacutainers (BD Biosciences) and processed the day of collection. Plasma was isolated and stored at −80°C.

### Cell Culture and Viruses

Vero E6 cells were obtained from ATCC (CRL-1586) and grown in DMEM supplemented with 10% heat-inactivated FBS, 2 mM L-glutamine, and 1x penicillin-streptomycin (cell culture medium). Cells were cultured under standard conditions and incubated at 37°C and 5% CO_2_.

MPXV clade IIb, lineage B.1, was obtained from BEI Resources (NR-58622). Viral stocks were prepared as previously described (10). Briefly, passage four viral stocks were grown at low multiplicity of infection (MOI = 0.01) in Vero E6 cells from sequence-verified passage three stocks. Following three days of viral growth, virus was harvested by freeze-thaw lysis of scraped cells followed by centrifugation at 2,000x*g*. Stocks were sucrose-purified using equal volumes of clarified supernatant and 36% sucrose in TNE buffer (Quality Biological #351-302-101) for ultracentrifugation at 32,900x*g* for 80 min at 4°C.

Virus was titered by immunofluorescent focus forming assay in Vero E6 cells. 1.5×10^4^ Vero E6 cells were plated in each well of a black 96-well assay plate in cell culture medium, then infected the following day with a 10-fold dilution curve of MPXV. Viral dilutions were prepared in DMEM lacking sodium pyruvate supplemented with 2% heat-inactivated FBS (MPXV infection media) and 5% guinea pig serum (Sigma-Aldrich G9774). Infection was allowed to progress for ~42 hours before a 1 h fixation at RT with 10% formalin (Fisher Scientific #SF984). Fixed samples were rinsed with water then permeabilized and blocked with 3% BSA in PBS (blocking buffer) with 0.1% Triton X-100 at RT for 30 min. Samples were then stained with a polyclonal rabbit anti-VACV Lister strain antibody (Abbexa abx023200) diluted 1:1,000 in blocking buffer for 1 h at RT. Plates were then washed four times with PBS before staining with a 1:2,000 dilution of donkey anti-rabbit AlexaFluor647 secondary (Thermo Scientific #A-31573) and 1.25 μg/mL DAPI in blocking buffer. Finally, cells were washed four times with PBS before filling each well with 100 μL of PBS and imaging on a BioTek Cytation 7 Cell Imaging Multi-Mode Reader with Gen5 Image Prime software for quantification. All MPXV work was conducted in a certified ABSL3 facility at NYU Grossman School of Medicine.

### Immunofluorecence-based MPXV Microneutralization Assay

MPXV neutralization assays were carried out using a modified version of the above MPXV focus forming assay. Briefly, on the day of infection, serum or plasma samples were diluted in MPXV infection media for a 7-point two-fold dilution curve starting at 1:20. All sera were previously complement-inactivated at 56°C for 30 min. These sera dilutions were mixed in equal volume with MPXV diluted in MPXV infection media with guinea pig serum (GPS). This mixing was accounted for to yield a final GPS concentration of 5%, an MOI of ~0.1, and an initial serum dilution of 1:20. Virus-serum mixtures were incubated for 1 h at 37°C and 5% CO_2_. Mixtures were then added to cells and incubated for 42 h. Fixation and subsequent staining steps were as above. MPXV neutralizing titers (ID_50_ values) were calculated from percent inhibition of infection using nonlinear regression (variable slope with four parameters) with top and bottom constraints (100 and 0, respectively) in GraphPad Prism version 10.4.2. All samples with ID_50_ values beyond the initial dilution range were repeated with a higher dilution series.

### Multiplexed Immunassay for Binding Antibodies and Avidity

Binding and avidity of OPXV-specific IgG in complement-inactivated serum or plasma were measured as previously described (10) with modifications detailed below. Briefly, ten different OPXV proteins were included in the panel: MPXV H3 (Sino Biological 40893-V08H1), MPXV A35 (Sino Biological 40886-V08H), MPXV A29 (Sino Biological 40891-V08E), MPXV A30 (Cell Sciences YVV15001A), MPXV M1 (Sino Biological 40904-V07H), MPXV B2 (Abbexa abx620100), MPXV A27 (Abbexa abx620105), MPXV B6 (Abbexa abx620118), VACV A33 (Sino Biological 40896-V07E), and VACV B5 (Sino Biological 40900-V08H). Three additional control beads were also included: human IgG1 (Sigma-Aldrich I5154), BSA (Fisher Scientific J65097.22), and IC45 (Luminex MRP1-045-01). Bead couplings were performed using the xMap Antibody Coupling Kit (Luminex 40-50016) with the following concentrations: 10 pmol/1×10^6^ beads for MPXV H3, MPXV A35, and human IgG1; 50 pmol/1×10^6^ beads for MPXV A29, MPXV A27, MPXV B6, VACV A33, and VACV B5; and 100 pmol/1×10^6^ beads for MPXV A30, MPXV M1, MPXV B2, and BSA. Coupling of OPXV proteins was confirmed using an anti-His antibody (Abcam ab27025) against the His-tagged antigens as well as a positive control pool of smallpox-vaccinated sera. Bead multiplexes were constructed using 1,000 beads per region for each sample.

Samples were analyzed by incubating equal parts serum or plasma and multiplexed beads for 1 h at RT in the dark on a shaker. Samples were analyzed at final dilutions of 1:400 and 1:3,200 prepared in duplicate in 1x PBS (−Ca^2+^ −Mg^2+^) with 0.01% BSA and 0.02% Tween-20 (PBS-TB). Beads were then washed twice with PBS-TB prior to incubation with either PBS (first replicate) or 2M ammonium thiocyanate (second replicate) for 30 min at RT in the dark on a shaker. The beads were washed again as before then incubated for 30 min as previously with biotinylated donkey anti-human IgG, Fcγ-specific (Jackson ImmunoResearch 709-065-098) diluted to 1 μg/mL in PBS-TB. Beads were washed once more and then incubated with streptavidin-PE (BioLegend 405204) diluted to 4 μg/mL in PBS-TB for 30 min as before. Following a final wash, beads were run on a Luminex 200 instrument with the following settings: sample volume set to 50 μL, 50 bead minimum per region, and DD gate set at 6,000 to 17,000. Binding titers were calculated by measuring area under the curve (AUC) of the median fluorescence intensity for the PBS condition. Avidity was calculated by taking the ratio of the AUCs of the ammonium thiocyanate condition over the PBS condition. Samples were normalized by the inclusion of a positive control pool on each plate, which was generated from pre-2022 control sera from 23 individuals with prior smallpox vaccination; normalization was conducted on an antigen-by-antigen basis. For all comparisons of avidity, only measurements from samples with above background binding were considered. Background levels of binding were measured for each antigen by use of 17 negative samples from pre-2022 with no prior smallpox vaccination. Threshold for positive binding was determined as two standard deviations above the mean AUC of negative samples.

### Serum CXCL13 Assay

Human serum CXCL13 was analyzed using the Simple Plex Human CXCL13 cartridge (ProteinSimple SPCKC-PS-000375) for the ELLA ProteinSimple instrument. Non-heat-inactivated sera samples were prepared following manufacturer’s instructions and diluted two-fold in kit diluent. For each sample, 50 μL of diluted serum was loaded and run on the ELLA ProteinSimple instrument.

### Statistical and Computational Methods

Statistical analyses were performed using the indicated test in GraphPad Prism version 10.4.2. Machine learning (ML) analysis was performed in Python using the scikit-learn (SKL) package version 0.24.2. For ML analyses, data were first scaled using the MinMaxScaler function from SKL on a column-by-column basis. Dimensionality reduction was performed using the SKL implementation of PCA to generate two PCs. The reduced dataset was then grouped by K means clustering based on silhouette scores (SS): SS of 0.548 for 2 clusters, SS of 0.549 for 3 clusters, SS of 0.540 for 4 clusters, and SS of 0.463 for 5 clusters. Using k = 4, samples from one experienced participant that were adjacent to the replicating-like cluster formed a separate cluster, but were grouped back with the replicating-like cluster for the purpose of analysis, yielding the three clusters detailed here. Classification of booster samples and samples from the Crandell et al. cohort was performed using the SKL implementation of a bagging classifier with a base support vector classifier. Bagging classifier used the following hyperparameters: gamma = 0.25, C = 128, 250 estimators, and max_samples = 0.9. Three pre-2022 naïve controls were excluded from the full analysis due to 1) elevated anti-MPXV A27 binding suggesting potential smallpox vaccination, 2) a significantly elevated MPXV neutralizing titer compared to other controls, and 3) above background levels of binding antibodies across four different antigens, respectively; this yields the 17 remaining negative controls detailed here. For PCA transformation and subsequent classification of Booster 1 samples, avidity measurements for anti-MPXV H3 were assayed using a higher dilution pair of 1:5,000 and 1:10,000 due to anti-MPXV H3 IgG titers measuring at the upper limit of quantitation following boosting. For all statistical tests, p < 0.05 is *, p < 0.01 is **, and p < 0.001 is ***.

### Study Approval

The NYC Observational Study of Mpox Immunity cohort has been registered with ClinicalTrials.gov (NCT05654883) and received approval from the NYU Institutional Review Board (protocol 22-01338). Additional samples obtained from the NYU Langone Vaccine Center Biorepository were under NYU Institutional Review Board protocol 18-02035. Samples from Crandell et al. (12) were obtained under the following authorities: Yale Human Research Protection Program Institutional Review Board (protocol ID 2000033415), the Research Ethics Committee of the Hospital Universitário Clementino Fraga Filho Universidade Federal do Rio de Janeiro (protocol number CAAE 62281722.5.0000.5257), and the Portuguese General Directorate of Health, through the technical orientation number 004/2022 on May 31, 2022. Written informed consent was obtained from all participants prior to participation in each study.

## Data Availability

All data produced in the present study are available upon reasonable request to the authors and will be fully available following publication.

## Author Contributions

**ALO**: Conceptualization, data curation, formal analysis, investigation, resources, methodology, project administration, validation, visualization, writing – original draft, and writing – review and editing. **KKW**: Investigation, methodology, writing – review and editing. **SR**: Data curation, project administration, resources, supervision, validation, and writing -- review and editing. **MT**: Resources, supervision, and writing – review and editing. **MIS**: Data curation, funding acquisition, project administration, supervision, and writing – review and editing. **ACK**: Project administration, supervision, and writing – review and editing. **RSH**: Supervision and writing – review and editing. **RD**: Data curation, supervision, validation, and writing – review and editing. **MJM**: Conceptualization, funding acquisition, project administration, supervision, and writing – review and editing.

## Acknowledgements

Most importantly, the authors thank the NYC OSMI study participants for their altruism and volunteerism, without whom we would have no study. The authors also wish to acknowledge the full NYC OSMI Study Group, which includes, in addition to the byline authors, the following colleagues: Abdul Abdulai; Robert Arciuolo, MPH, CPH; Jaqueline Callahan, RN; Ellie Carmody, MD, MPH; Tamia Davis, NP; Amanda Dontino; Aimee Edwin, RN; Celia Engelson, NP; Andrew Fleming, MD; Olivia Frank, MPH; Emily Geesey; Shelby Goins; Victoria Guerra, RN; Erika Guevarra; Sarah Haiken; Aaliyah Henry; Kathryn Jano; Trishala Karmacharya; Dorothy Knutsen, MD; Irma Noriega, NP; Maria Null; Samuel Nweke; Lalitha Parameswaran, MD, MPH; Robert Pitts, MD, MPH; Gurchetan Randhawa, MD; Miguel Rodriguez; Pamela Suman; Apoorva Talanki; Meron Tasissa; Kathryn Swindell, NP; Julia Wagner, MPH; Jimmy P. Wilson; Samantha Yip, RN; Miilani Yonatan; Heekoung Allison Youn, RN; Lisa Zhao. The authors acknowledge the generous sharing of samples and feedback from Dr. Carolina Lucas and Jay Crandell. The authors want to thank Drs. Jane Zucker and Jennifer Rosen from the NYC Department of Health and Mental Hygiene for their support in obtaining funding for this study. We also thank the NYU Office of Scientific Research Animal Biosafety Level 3 staff, Drs. Ludovic Desvignes, Dominick Papandrea, and Alison Gilchrist, for their support of this work. Financial support was provided by the Blavatnik Family Foundation, the National Institutes of Health (NIH) grants no. AI148574 and 75N93021C00014 (to M.J.M.) and T32AI100853 (supporting A.L.O.), the NYC Department of Health and Mental Hygiene, and NYU Grossman School of Medicine institutional support.

