## Supplemental Materials for "A three-dose MVA-BN mpox vaccination series improves the quality of anti-monkeypox virus immunity"

**Supplementary Table S1:** Demographics and patient data for MPXV convalescent participants.

| Subject | Age Range | Gender | HIV status | CD4 count | Received Tecovirimat? | Vax #1 route | Vax #2 route | Smallpox Vax | Vax 1 DPO | Vax 2 DPO | Year 1 Cluster | Year 2 Cluster |
|---|---|---|---|---|---|---|---|---|---|---|---|---|
| Conv 1 | 60-69 | Male | HIV positive | 640 | No | | | Yes | | | RL | RL |
| Conv 2 | 30-39 | Male | HIV positive | 526 | Yes | | | No | | | RL | RL |
| Conv 3 | 30-39 | Male | HIV negative | | No | SC | ID | No | 9 | 87 | NRL | NRL |
| Conv 4 | 50-59 | Male | HIV positive | 1100 | Yes | | | Yes | | | RL | RL |
| Conv 5 | 40-49 | Male | HIV positive | 553 | Yes | SC | ID | No | -25 | 46 | NRL | NRL |
| Conv 6 | 20-29 | Male | HIV negative | | Yes | SC | | No | -1 | | NRL | |
| Conv 7 | 30-39 | Male | HIV positive | 730 | No | | | No | | | NRL | NRL |
| Conv 8 | 40-49 | Male | HIV positive | 1158 | Yes | ID | | No | 78 | | NRL | RL |
| Conv 9 | 60-69 | Male | HIV positive | 928 | No | | | Yes | | | RL | RL |

### Supplementary Figure S1

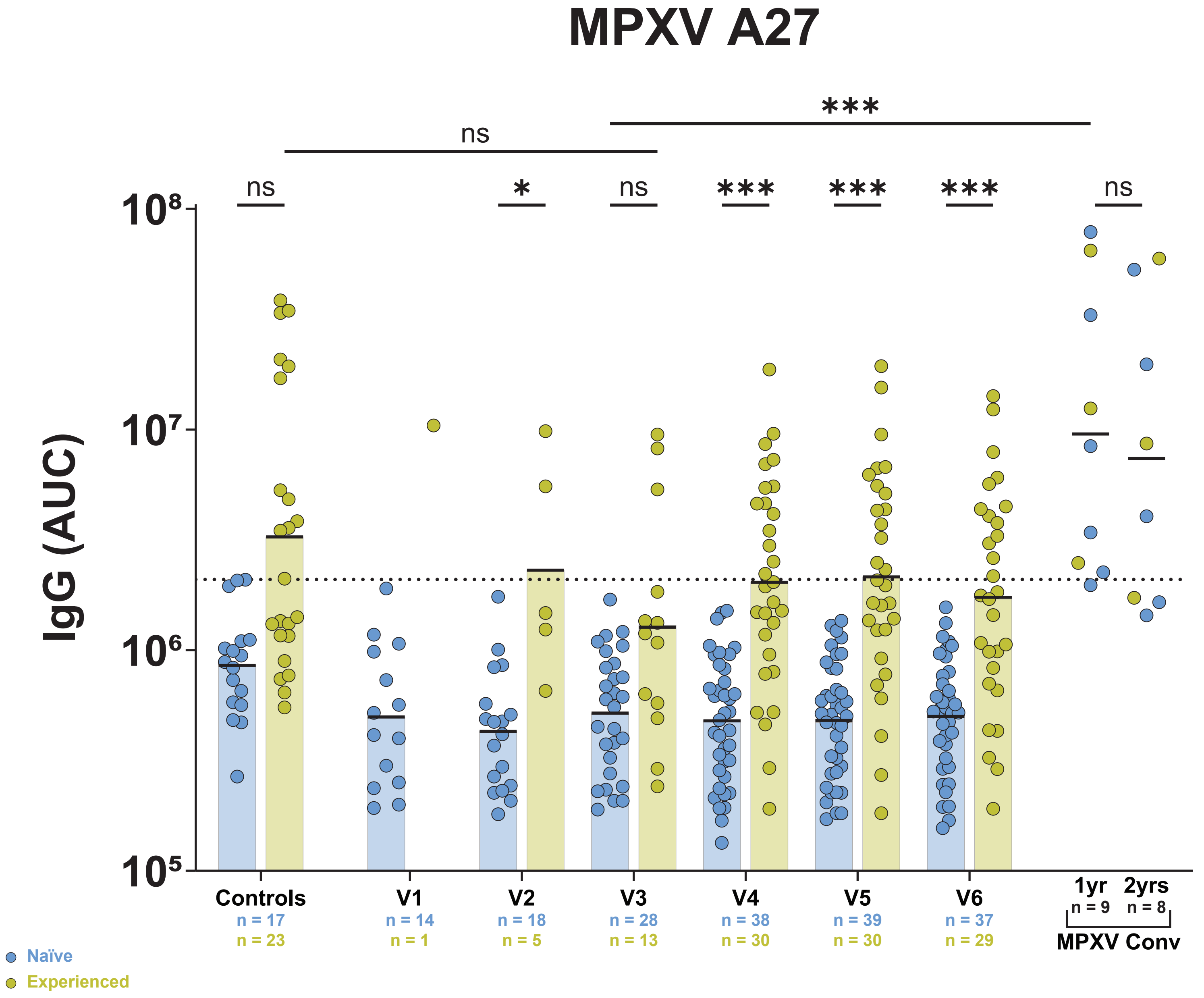

**Supplementary Figure S1: Above background levels of anti-MPXV A27 are found only among participants with history of replicating OPXV exposure.** Anti-MPXV A27 IgG were assayed using multiplexed immunoassay. Dashed line represents positivity threshold as determined by two times the standard deviation of the pre-2022 naïve controls plus their average. Black bars represent the geometric mean. Statistics were performed using the Kruskal-Wallis test with Dunn’s method for multiple comparisons. MPXV Conv, mpox convalescent patients.

### Supplementary Figure S2

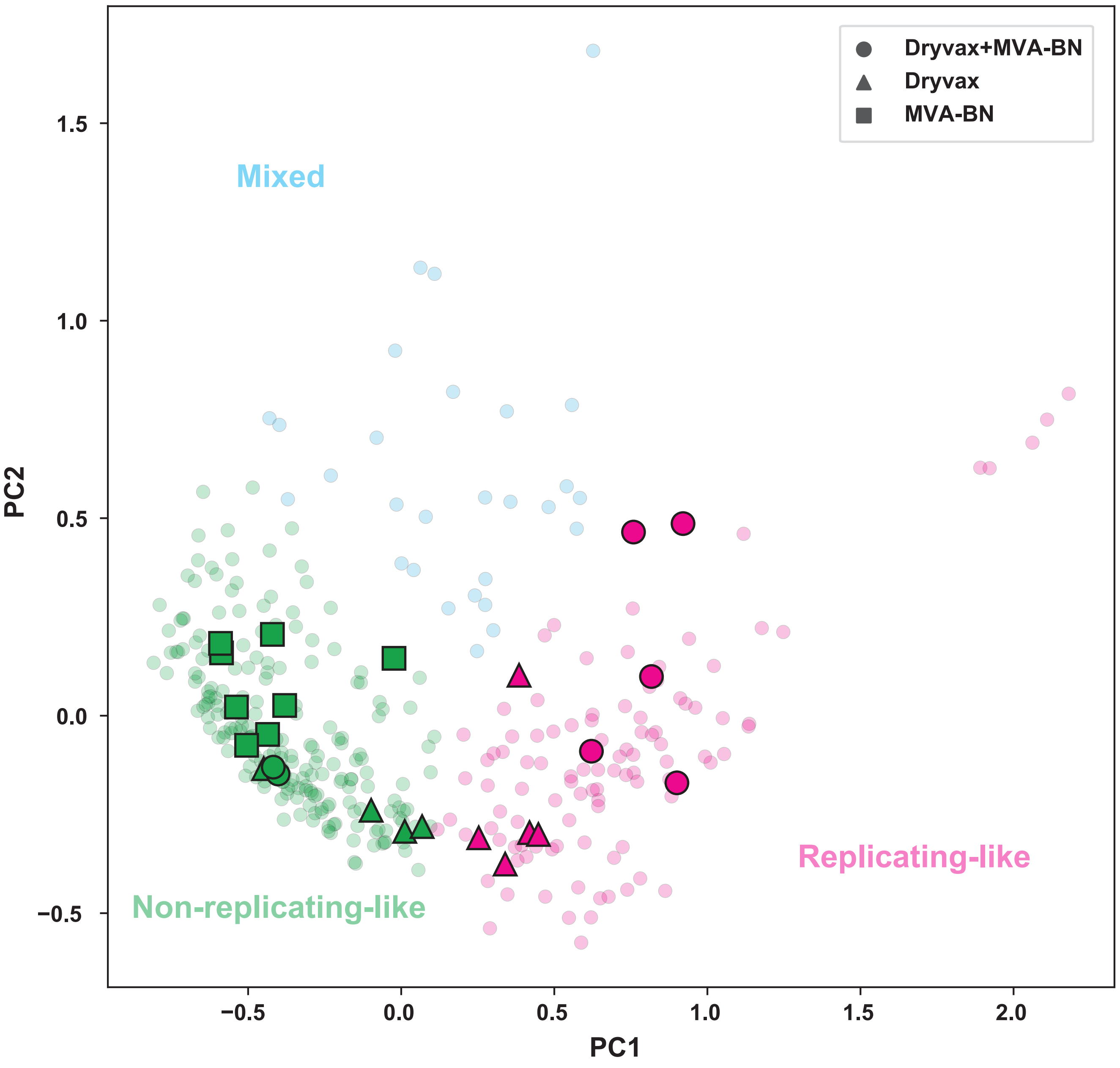

**Supplementary Figure S2: Samples from the cohort described in Crandell et al. (12) behave similarly to NYC OSMI samples.** PCA plot of Crandell et al. samples colored according to classification using a bagged support vector machine classifier trained on NYC OSMI samples.
